# Pre-pandemic psychiatric disorders and risk of COVID-19: a cohort analysis in the UK Biobank

**DOI:** 10.1101/2020.08.07.20169847

**Authors:** Huazhen Yang, Wenwen Chen, Yao Hu, Yilong Chen, Yu Zeng, Yajing Sun, Zhiye Ying, Junhui He, Yuanyuan Qu, Donghao Lu, Fang Fang, Unnur A Valdimarsdóttir, Huan Song

## Abstract

**Objective:** To determine the association between pre-pandemic psychiatric disorders and the risk of COVID-19.

**Design:** Community-based prospective cohort study.

**Setting:** UK Biobank population.

**Participants:** 421,048 participants who were recruited in England and alive by January 31^st^ 2020, i.e., the start of COVID-19 outbreak in the UK. 50,815 individuals with psychiatric disorders recorded in the UK Biobank inpatient hospital data before the outbreak were included in the exposed group, while 370,233 participants without such conditions were in the unexposed group.

**Measurements:** We obtained information on positive results of COVID-19 test as registered in the Public Health England, COVID-19 related hospitalizations in the UK Biobank inpatient hospital data, and COIVD-19 related deaths from the death registers. We also identified individuals who was hospitalized for infections other than COVID-19 during the follow-up. Logistic regression models were used to estimate odds ratios (ORs) with 95% confidence intervals (CIs), controlling for multiple confounders.

**Results:** The mean age at outbreak was 67.8 years and around 43% of the study participants were male. We observed an elevated risk of COVID-19 among individuals with pre-pandemic psychiatric disorder, compared with those without such diagnoses. The fully adjusted ORs were 1.44 (95%CI 1.27 to 1.64), 1.67 (1.42 to 1.98), and 2.03 (1.56 to 2.63) for any COVID-19, inpatient COVID-19, COVID-19 related death, respectively. The excess risk was observed across all levels of somatic comorbidities and subtypes of pre-pandemic psychiatric disorders, while further increased with greater number of pre-pandemic psychiatric disorders. We also observed an association between pre-pandemic psychiatric disorders and increased risk of hospitalization for other infections (1.85 [1.65 to 2.07]).

**Conclusions:** Pre-pandemic psychiatric disorders are associated with increased risk of COVID-19, especially severe and fatal COVID-19. The similar association observed for hospitalization for other infections suggests a shared pathway between psychiatric disorders and different infections, including altered immune responses.

**Summary box:** *What is already known on this topic?:* Psychiatric morbidities have been associated with risks of severe infections through compromised immunity and/or health-behaviors. While recent studies showed that unhealthy lifestyle and psychosocial factors (including self-reported psychological distress) increased the risk of COVID-19 hospitalization, data on the role of clinically confirmed psychiatric disorders in COVID-19 susceptibility are to date absent.

*What this study adds?:* Using the large community-based data in UK Biobank, our analysis is the first to demonstrate an increase in the risk of COVID-19, especially severe and fatal COVID-19, among individuals with pre-pandemic psychiatric disorders, independently of many important confounders. A similar association was also observed between pre-pandemic psychiatric disorder and hospitalization due to other infections during the COVID-19 outbreak, suggesting a shared pathway between psychiatric disorders and different infections, including altered immune responses. This finding underscores the need of surveillance and care in vulnerable populations with history of psychiatric disorders during the COVID-19 outbreak.

## Introduction

The Coronavirus disease 2019 (COVID-19) pandemic, caused by severe acute respiratory syndrome coronavirus 2 (SARS-CoV-2), is posing an unprecedented crisis worldwide^1^. Given its rapid transmission and the considerable proportion of severe cases needing intensive medical care, this new and potent infectious disease has brought a major pressure on the global healthcare system and the general public. According to the World Health Organization (WHO), the SARS-CoV-2 has spread to over 100 countries, infected over 17.9 million people, and led to more than 686,000 deaths as of August 3^rd^, 2020^2^, and the number keeps increasing.

Psychiatric morbidities, such as depression, anxiety, and stress-related disorders, have consistently been associated with an elevated risk of a number of somatic diseases including autoimmune disease^3^, respiratory diseases^4^, and severe infections^5 6^, possibility through altered immune responses^7^. It is therefore plausible that prior psychiatric disorders may also alter individual susceptibility to COVID-19. Emerging evidence has shown a strong genetic correlation between having at least one psychiatric diagnosis and the occurrence of infection and lends further support to this notion^8^. It is further possible that suboptimal behaviors (e.g., smoking), as well as incapability for mitigation strategies during the COVID-19 outbreak, among individuals with psychiatric disorders can contribute to an increased vulnerability to COVID-19. Indeed, recent studies showed that unhealthy lifestyle^9^ and psychosocial factors (including self-reported psychological distress)^10^ increased the risk of COVID-19 hospitalization. Yet, data on the role of clinically confirmed psychiatric disorders in COVID-19 susceptibility from longitudinal study are to date absent.

Taking advantage of the rich information on phenotypes relevant to mental health, other life-style factors and the continuously updated data on COVID-19 infection in the UK Biobank, we aimed to determine the association between pre-pandemic psychiatric disorders and the subsequent risk of COVID-19.

## Methods

### Study design

UK Biobank is a prospective cohort study of 502,507 middle-aged (40-69 years old) participants across England, Scotland and Wales between 2006 and 2010. Information about socio-demographic, lifestyle and health-related factors was collected at recruitment^11^. Health-related outcomes were obtained through periodically linked data from multiple national datasets, with participants’ consent^12^. The inpatient hospital data were mapped across England, Scotland and Wales based on Hospital Episode Statistics in England, Scottish Morbidity Record, and Patient Episode Database for Wales^13^. Data on mortality are updated from NHS Digital for participants in England and Wales and from the NHS Central Register for participants in Scotland^14^. After the global outbreak of COVID-19, UK Biobank has been additionally linked to Public Health England (PHE) where the results of COVID-19 tests, performed by real-time RT-PCR (RdRp gene assay) based on oral swabs, in England were released from March 16^th^ 2020 onward^15 16^.

The present analysis is limited participants who registered in England (n=445,858) as no data on COVID-19 tests were available for participants in Scotland and Wales. We also excluded 24,810 individuals who died before January 31^st^, 2020, when the first COVID-19 case was diagnosed in the UK, leaving 421,048 participants for further analysis (shown in Figure 1). Individuals with a diagnosis of psychiatric disorders before the date of COVID-19 outbreak were considered having a pre-pandemic psychiatric disorder and included in the exposed group, while the others were included in the unexposed group.

**Figure 1.**
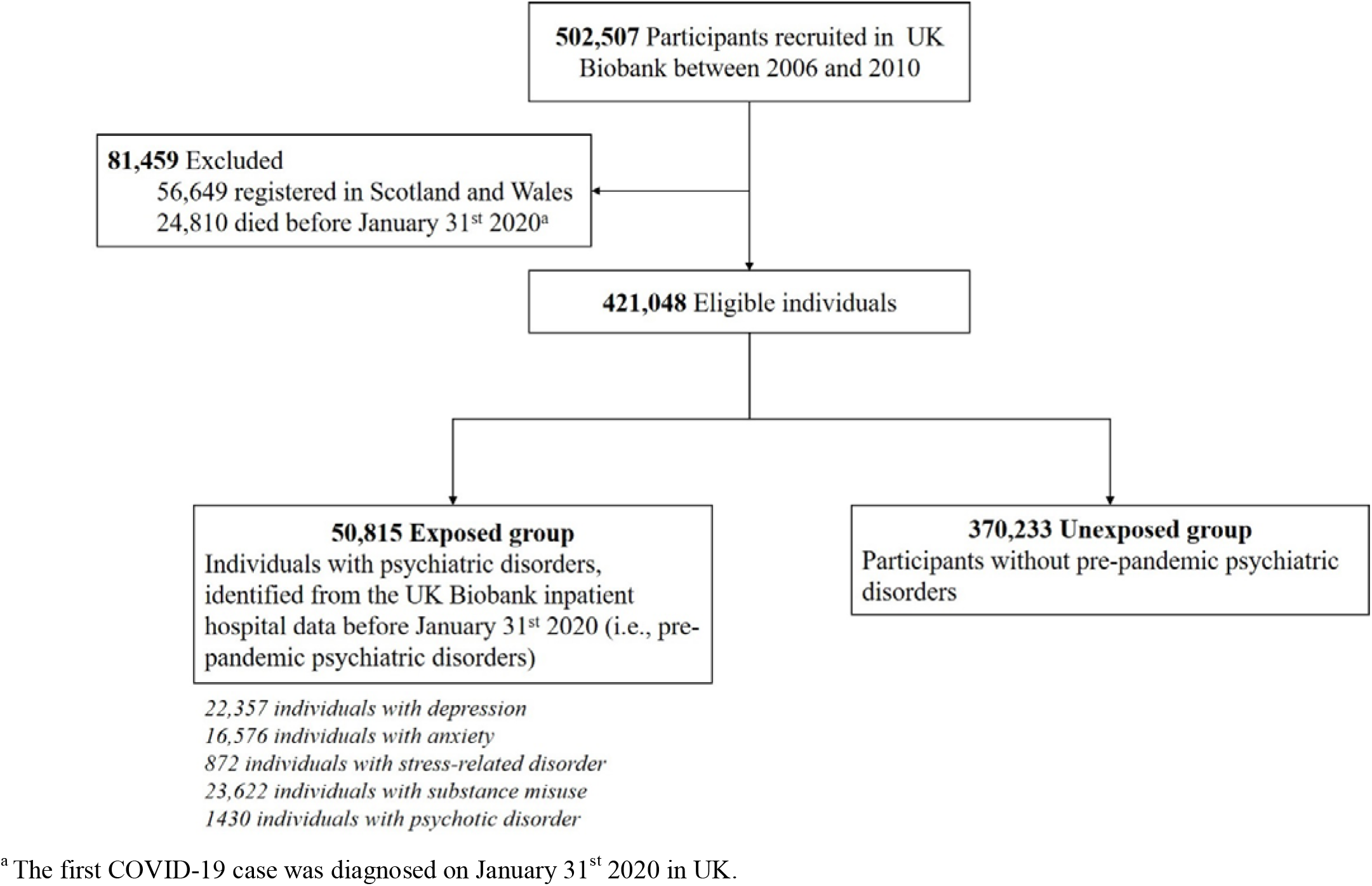
Study design ^a^The first COVID-19 case was diagnosed on January 31st 2020 in UK.

All UK Biobank participants gave written informed consent prior to the data collection. UK Biobank has full ethical approval from the NHS National Research Ethics Service (reference number: 16/NW/0274), and the study was also approved by the biomedical research ethics committee of West China Hospital (reference number: 2020.661).

#### Pre-pandemic psychiatric disorders

We retrived information about the diagnoses of psychiatric disorders from the UK Biobank inpatient hospital data, available since 1981. A pre-pandemic psychiatric disorder was defined as any hospital admission with a diagnosis of psychiatric disorders, including depression (according to the International Classification of Diseases [ICD]-10 codes: F32-F33, ICD-9: 296.1, 300.4 and 311), anxiety (ICD-10: F40-F41, ICD-9: 300.0 and 300.2), stress-related disorders (ICD-10: F43, ICD-9: 308 and 309), substance misuse (ICD-10: F10-F19, ICD-9: 291, and 303-305), and psychotic disorders (ICD-10: F20-F29, ICD-9: 295, 297 and 298), before January 31^st^ 2020.

#### Ascertainment of COVID-19

The ascertainment of COVID-19 mainly relied on the PHE dataset, which contains information about specimen date, origin (inpatient or not), and result (positive or negative) of all COVID-19 tests performed in England between March 16^th^ and July 19^th^ 2020^16^. Further, we additionally used the UK Biobank inpatient hospital data and death registers, i.e., any diagnosis or death cause coded as U07.1 and U07.2 (ICD-10), to ensure the completement of COVID-19 case identification. The caseness acertained by any of the above-mentioned approaches was mentioned as ‘all COVID-19’.

Notably, the testing strategy varied over time in the UK. The initial tests were largely restricted to inpatients with symptoms, and a positive result was therefore a reasonable proxy for severe COVID-19. As the tesing capacity increased, after 27^th^ April, more community testing was introduced and all patients admitted to hospital for overnight, including asymptomatic ones, received the test^15^. Accordingly, besides all COVID-19, we conducted analyses specificially for inpatient COVID-19, defined a hospital admission with a diagnosis of COVID-19 according to the UK Biobank inpatient hospital data, or a positive test from PHE provided that the origin of test was marked as ‘inpatient’.

We defined COVID-19-related death as a death with COVID-19 (ICD-10: U07.1 and U07.2) as the underlying cause according to data from the death registers (updated until 22^nd^ May 2020).

#### Hopitalization for other infections

To test our hypothesis that individuals with pre-pandemic psychiatric disorders are actually more vulnerable to all severe infections^5^, including COVID-19, through for instance compromised immunity, we used other infections that required hospital care during the study period as a positive disease control. Individuals hospitalized for other infections were defined as the ones who were free of COVID-19 and admitted to hospital with a main diagnosis of other infections (ICD codes in Supplementary Table 1) between January 31^st^ and March 31^st^ 2020, based on the UK Biobank inpatient hospital data.

#### Covariates

Data on sociodemographic (e.g., birth year, sex, and ethnicity) and lifestyle factors (e.g., smoking status) were collected at baseline through questionnaires. Body mass index (BMI) was constructed from height and weight measured during the initial assessment center visit. The postcode of study participants was used to generate the Townsend deprivation index, which is widely used as a measure of area-level socioeconomic deprivation, with higher scores representing greater deprivation^17^. In addition, history of somatic diseases that could affect the COVID-19 susceptibility or course (i.e., chronic cardiac disease, diabetes, chronic pulmonary disease, chronic kidney disease, and asthma, with ICD codes listed in Supplementary Table1) were selected according to previous literature^18-20^ and were also extracted from the UK Biobank inpatient hospital data.

### Statistical Analysis

We examined the association between pre-pandemic psychiatric disorders and risk of COVID-19 using odds ratios (ORs) with 95% confidence intervals (CIs) using logistic regression models. We first performed analyses for all COVID-19, and then specifically for inpatient COVID-19 and COVID-19 related death. All models were adjusted for birth year, sex (male or female), race/ethnicity (White, others, or unknown), BMI level (<18.5 kg/m^2^, 18.5-24.9 kg/m^2^, 25.0-29.9 kg/m^2^, or ≥30 kg/m^2^), smoking status (never, ever, or prefer not to answer), Townsend deprivation index (as continuous variable), and the history of chronic cardiac disease (yes or no), diabetes (yes or no), chronic pulmonary disease (yes or no), chronic kidney disease (yes or no), and asthma (yes or no).

In subgroup analyses, we calculated the ORs by sex (male or female), age at the outbreak (i.e., on 31^st^ Jan 2020, by tertiles, ≤64, 65-72, or ≥73 years), BMI level (<18.5 kg/m2, 18.5-24.9 kg/m2, 25.0-29.9 kg/m2, or ≥30 kg/m2), smoking status (never or ever), the number of somatic comorbidities (0, 1, 2, or ≥3), and Townsend deprivation score (by tertiles, low[≤-3.15], middle [-3.16~-0.63], or high [≥-0.64]). Further, to explore whether the impact of pre-pandemic psychiatric disorders on COVID-19 susceptibility varied during different stages of the outbreak, we assessed the studied association by different time periods (before March 31^st^, April 1^st^ to May 31^st^, or June 1^st^ 2020 onward).

Additionally, we also separately investigated the association according to the time since the first diagnosis of psychiatric disorders (<1, 1-2, 3-4, 5-9, or ≥10 years) and the number of pre-pandemic psychiatric disorders (1, 2, or ≥3). In addition to considering all pre-pandemic psychiatric disorders as one group, we did separate analyses for five subtypes of psychiatric disorders, i.e., depression, anxiety, stress-related disorder, substance misuse, and psychotic disorder.

To explore the impact of pre-pandemic psychiatric disorders on susceptibility of other severe infections, excluding COVID-19, we re-ran all these analyses for hospitalization for other infections. Next, we repeated all main analyses by using the primary diagnoses of psychiatric disorders only in the UK Biobank inpatient hospital data, to test the robustness of the results to the definition of pre-pandemic psychiatric disorders. Moreover, because the testing strategy in UK changed considerably on 27^th^ April, we did another sensitivity analysis by restricting to the study period before this date. All analyses were conducted in R software, version 3.6. A 2-sided *p* < 0.05 was considered statistically significant.

### Patient and public involvement

No patients were engaged in setting the research question or the outcome measures, nor were they involved in design or implementation of the study. There are no plans to disseminate the results of the research directly to study participants or the relevant patient community. Results from UK Biobank are routinely disseminated to study participants via the study website and social media outlets.

## Results

We in total identified 50,815 individuals with pre-pandemic psychiatric disorder (12.1% of all eligible participants), and the number of unposed individuals was 370,233 (87.9%). The mean age at the time of COVID-19 outbreak was 67.8 years and around 43% of the study participants were male (Table 1). While there was no difference with regard to age, sex, and race, individuals with pre-pandemic psychiatric disorders were more likely to be smokers (63.9% vs 41.4%), with higher BMI (≥30 kg/m^2^: 31.8% vs 23.3%) and greater socioeconomic deprivation (i.e., Townsend deprivation index: −0.38 vs −1.46), compared with the unexposed individuals. We also found higher proportions of exposed individuals to have a history of severe somatic comorbidities, including chronic cardiac disease (21.7% vs 7.8%), diabetes (14.9% vs 6.3%), chronic pulmonary disease (14.3% vs 3.3%), chronic kidney disease (7.0% vs 2.7%) and asthma (17.4% v 7.9%).

**Table 1.**
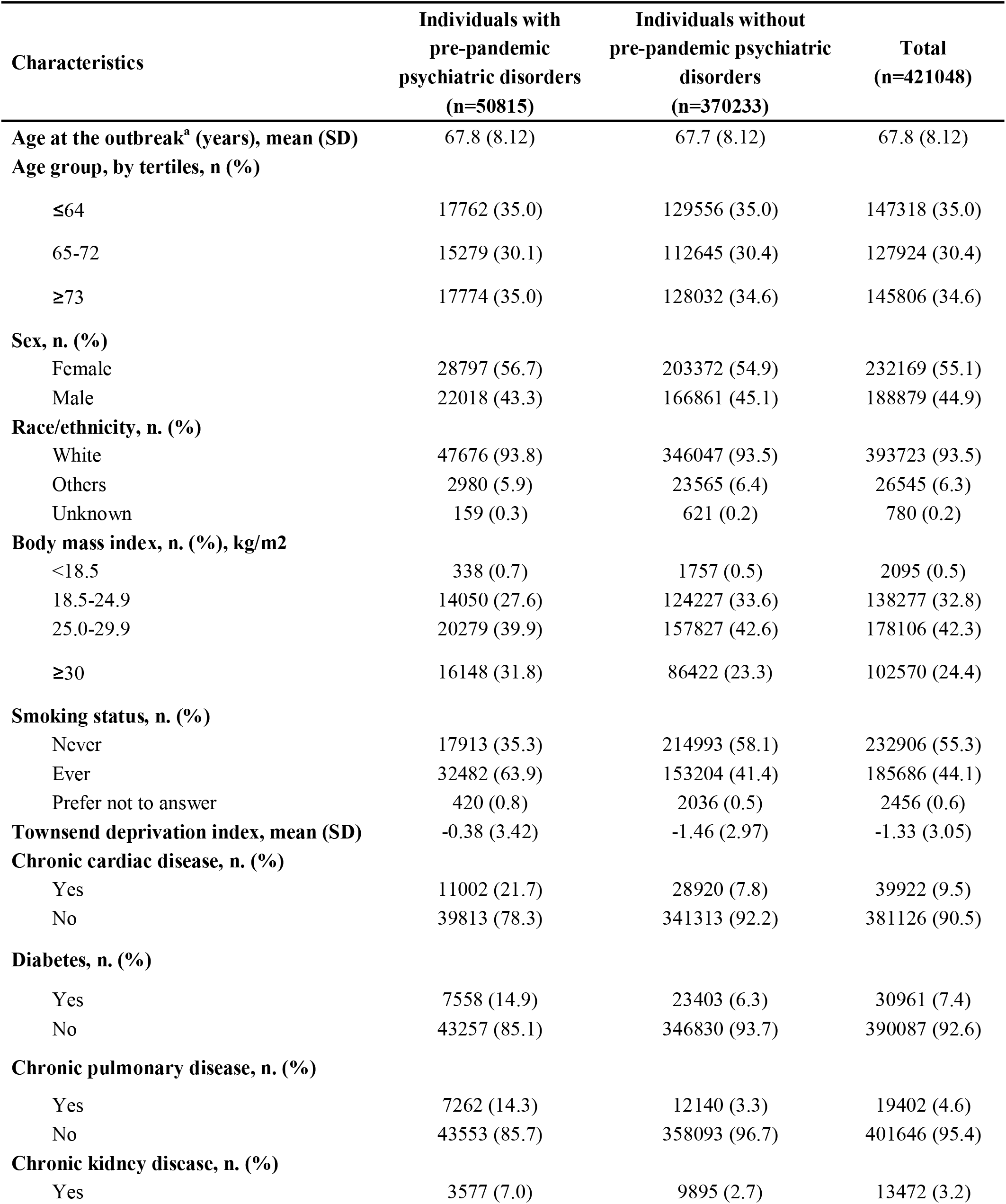
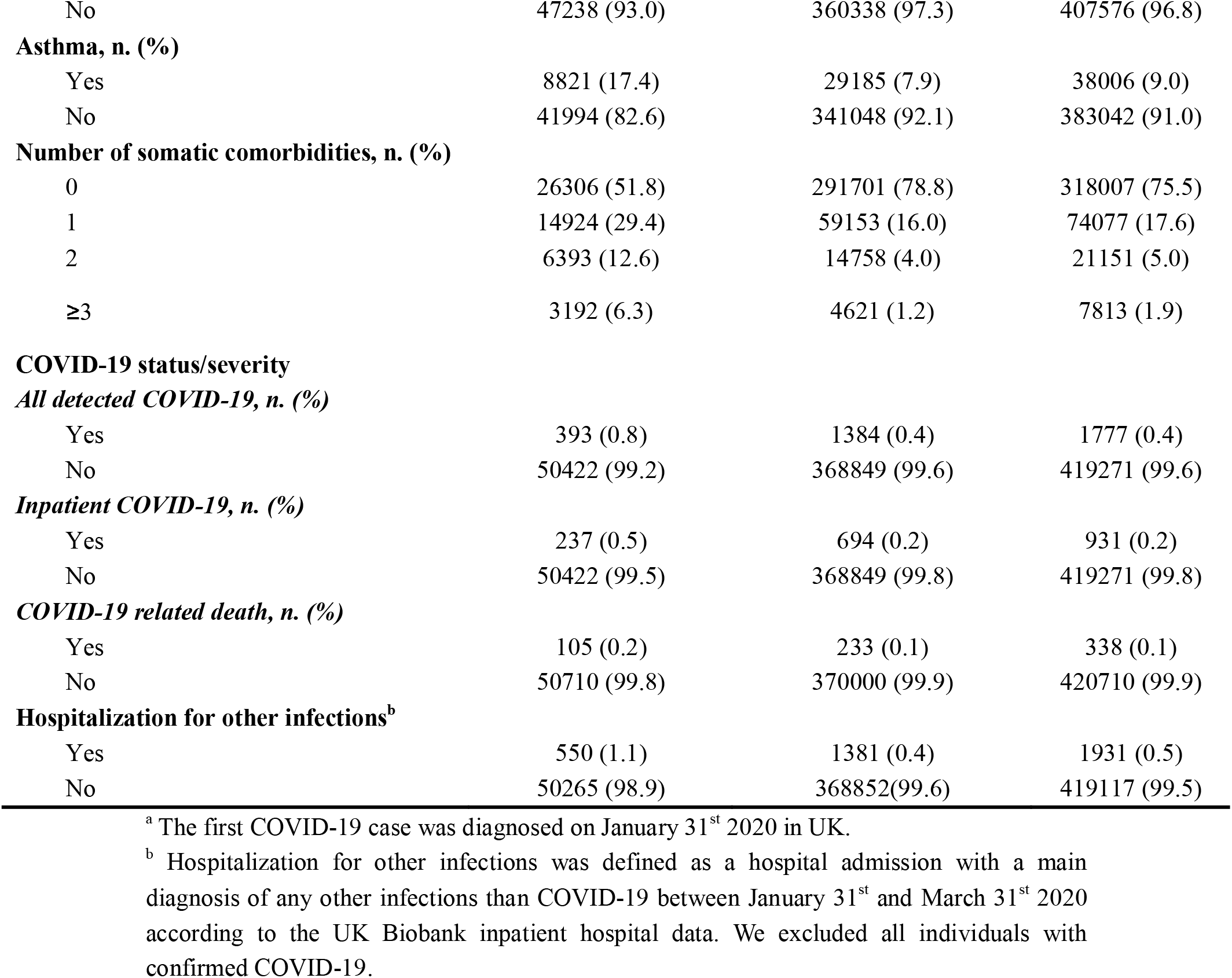
Characteristics of the study cohort

| Characteristics | Individuals with<br>pre-pandemic<br>psychiatric disorders<br>(n=50815) | Individuals without<br>pre-pandemic psychiatric<br>disorders<br>(n=370233) | Total<br>(n=421048) |
| --- | --- | --- | --- |
| **Age at the outbreak<sup>a</sup> (years), mean (SD)** | 67.8 (8.12) | 67.7 (8.12) | 67.8 (8.12) |
| **Age group, by tertiles, n (%)** |  |  |  |
| ≤64 | 17762 (35.0) | 129556 (35.0) | 147318 (35.0) |
| 65-72 | 15279 (30.1) | 112645 (30.4) | 127924 (30.4) |
| ≥73 | 17774 (35.0) | 128032 (34.6) | 145806 (34.6) |
| **Sex, n. (%)** |  |  |  |
| Female | 28797 (56.7) | 203372 (54.9) | 232169 (55.1) |
| Male | 22018 (43.3) | 166861 (45.1) | 188879 (44.9) |
| **Race/ethnicity, n. (%)** |  |  |  |
| White | 47676 (93.8) | 346047 (93.5) | 393723 (93.5) |
| Others | 2980 (5.9) | 23565 (6.4) | 26545 (6.3) |
| Unknown | 159 (0.3) | 621 (0.2) | 780 (0.2) |
| **Body mass index, n. (%), kg/m<sup>2</sup>** |  |  |  |
| <18.5 | 338 (0.7) | 1757 (0.5) | 2095 (0.5) |
| 18.5-24.9 | 14050 (27.6) | 124227 (33.6) | 138277 (32.8) |
| 25.0-29.9 | 20279 (39.9) | 157827 (42.6) | 178106 (42.3) |
| ≥30 | 16148 (31.8) | 86422 (23.3) | 102570 (24.4) |
| **Smoking status, n. (%)** |  |  |  |
| Never | 17913 (35.3) | 214993 (58.1) | 232906 (55.3) |
| Ever | 32482 (63.9) | 153204 (41.4) | 185686 (44.1) |
| Prefer not to answer | 420 (0.8) | 2036 (0.5) | 2456 (0.6) |
| **Townsend deprivation index, mean (SD)** | -0.38 (3.42) | -1.46 (2.97) | -1.33 (3.05) |
| **Chronic cardiac disease, n. (%)** |  |  |  |
| Yes | 11002 (21.7) | 28920 (7.8) | 39922 (9.5) |
| No | 39813 (78.3) | 341313 (92.2) | 381126 (90.5) |
| **Diabetes, n. (%)** |  |  |  |
| Yes | 7558 (14.9) | 23403 (6.3) | 30961 (7.4) |
| No | 43257 (85.1) | 346830 (93.7) | 390087 (92.6) |
| **Chronic pulmonary disease, n. (%)** |  |  |  |
| Yes | 7262 (14.3) | 12140 (3.3) | 19402 (4.6) |
| No | 43553 (85.7) | 358093 (96.7) | 401646 (95.4) |
| **Chronic kidney disease, n. (%)** |  |  |  |
| Yes | 3577 (7.0) | 9895 (2.7) | 13472 (3.2) |
| No | 47238 (93.0) | 360338 (97.3) | 407576 (96.8) |
| **Asthma, n. (%)** |  |  |  |
| Yes | 8821 (17.4) | 29185 (7.9) | 38006 (9.0) |
| No | 41994 (82.6) | 341048 (92.1) | 383042 (91.0) |
| **Number of somatic comorbidities, n. (%)** |  |  |  |
| 0 | 26306 (51.8) | 291701 (78.8) | 318007 (75.5) |
| 1 | 14924 (29.4) | 59153 (16.0) | 74077 (17.6) |
| 2 | 6393 (12.6) | 14758 (4.0) | 21151 (5.0) |
| ≥3 | 3192 (6.3) | 4621 (1.2) | 7813 (1.9) |
| **COVID-19 status/severity** |  |  |  |
| ***All detected COVID-19, n. (%)*** |  |  |  |
| Yes | 393 (0.8) | 1384 (0.4) | 1777 (0.4) |
| No | 50422 (99.2) | 368849 (99.6) | 419271 (99.6) |
| ***Inpatient COVID-19, n. (%)*** |  |  |  |
| Yes | 237 (0.5) | 694 (0.2) | 931 (0.2) |
| No | 50422 (99.5) | 368849 (99.8) | 419271 (99.8) |
| ***COVID-19 related death, n. (%)*** |  |  |  |
| Yes | 105 (0.2) | 233 (0.1) | 338 (0.1) |
| No | 50710 (99.8) | 370000 (99.9) | 420710 (99.9) |
| **Hospitalization for other infections<sup>b</sup>** |  |  |  |
| Yes | 550 (1.1) | 1381 (0.4) | 1931 (0.5) |
| No | 50265 (98.9) | 368852(99.6) | 419117 (99.5) |

<sup>a</sup> The first COVID-19 case was diagnosed on January 31<sup>st</sup> 2020 in UK.
<sup>b</sup> Hospitalization for other infections was defined as a hospital admission with a main diagnosis of any other infections than COVID-19 between January 31<sup>st</sup> and March 31<sup>st</sup> 2020 according to the UK Biobank inpatient hospital data. We excluded all individuals with confirmed COVID-19.

As of July 19^th^, we identified a total of 1,777 COVID-19 cases, among which 931 were inpatient COVID-19 cases. The number of COVID-19 related deaths until 22^nd^ May was 338 (Table 1). In general, we observed elevated risk of COVID-19 among individuals with pre-pandemic psychiatric disorders, compared with individuals without such a condition (Figure 2). The proportion of all COVID-19, inpatient COVID-19, and COVID-19 related death was 0.8%, 0.5%, and 0.2%, respectively in the exposed group, and was 0.4%, 0.2%, and 0.1%, respectively, in the unexposed group. This corresponded to an adjusted relative risk of 1.44 (95%CI 1.27 to 1.64) for all COVID to 19. The fully adjusted OR was 1.67 (95%CI 1.42 to 1.98) for inpatient COVID to 19 and 2.03 (95%CI 1.56 to 2.63) for COVID-19 related death (Figure 2). The observed associations were largely comparable between all studied subtypes of pre-pandemic psychiatric disorders, with the highest point estimate observed for psychotic disorder, followed by stress-related disorder and depression (Figure 2). Similar result patterns were observed for hospitalization for other infections (Figure 2). The fully adjusted OR of hospitalization for other infections was 1.85 (95% CI 1.65 to 2.07) for any pre-pandemic psychiatric disorder, and 2.50 (95%CI 1.68 to 3.72) for pre-pandemic psychotic disorder.

**Figure 2.**
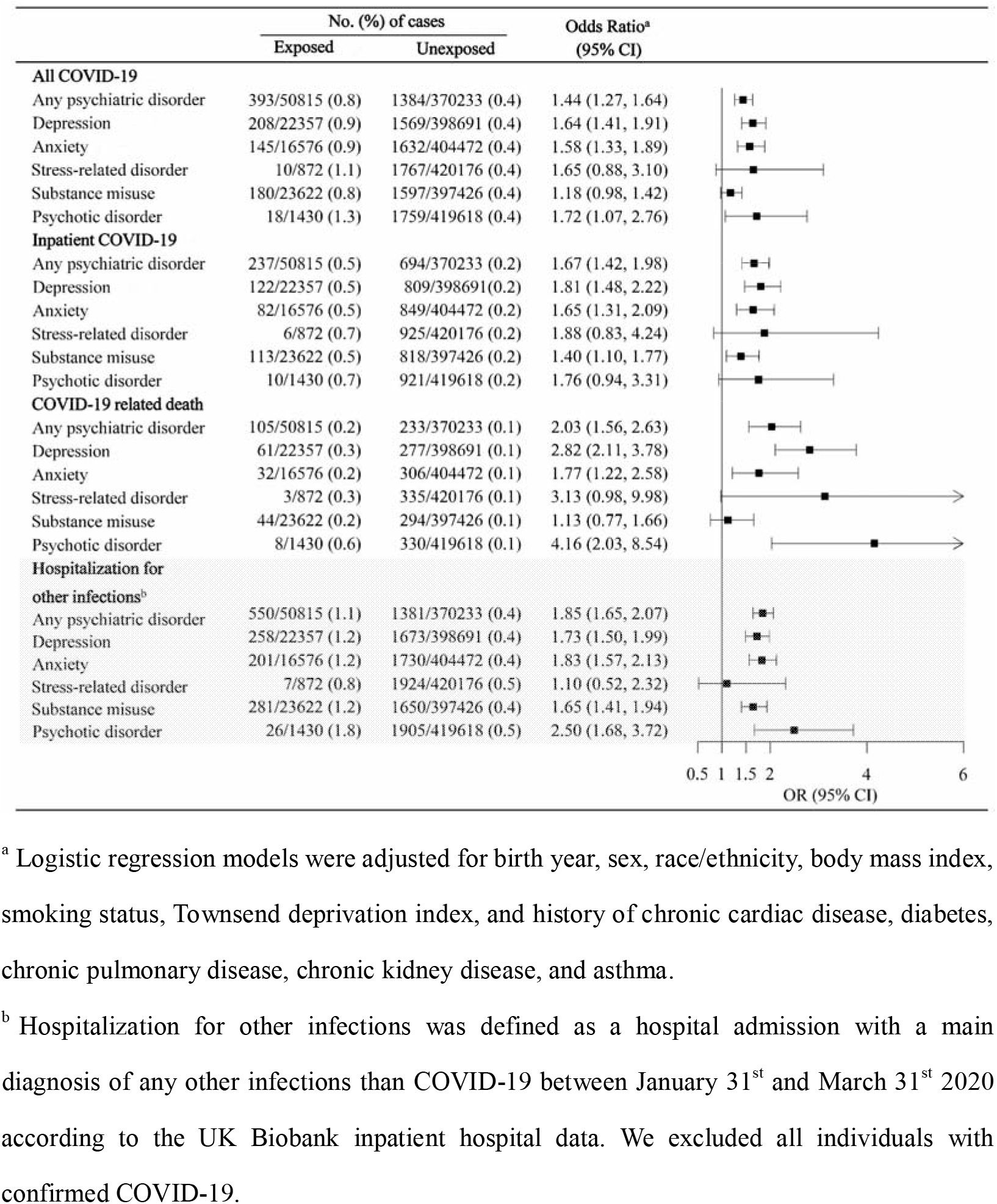
Risk of COVID-19 and other infections among individuals with any or specific pre-pandemic psychiatric disorder, compared to individuals without such a condition ^a^Logistic regression models were adjusted for birth year, sex, race/ethnicity, body mass index, smoking status, Townsend deprivation index, and history of chronic cardiac disease, diabetes, chronic pulmonary disease, chronic kidney disease, and asthma. ^b^Hospitalization for other infections was defined as a hospital admission with a main diagnosis of any other infections than COVID-19 between January 31st and March 31st 2020 according to the UK Biobank inpatient hospital data. We excluded all individuals with confirmed COVID-19.

The associations between pre-pandemic psychiatric disorders and COVID-19 did not differ by sex, BMI level, smoking status, the number of somatic comorbidities, level of Townsend deprivation score, or the stage of COVID-19 outbreak (Table 2). However, the associations appeared to be stronger among individuals older than 64 years, for all COVID-19 (OR was 0.85 [95%CI 0.66 to 1.10], 1.81 [95%CI 1.39 to 2.35], and 1.83 [95%CI 1.54 to 2.19] for individuals aged ≤64, 65-72, and ≥73 years, respectively).

**Table 2.** Risk of COVID-19 among individuals with pre-pandemic psychiatric disorders, by different characteristics, compared to individuals without such

|  | No. (%) of all COVID-19 |  | Odds Ratio<sup>a</sup><br>(95% CI) | No. (%) of inpatient COVID-19 |  | Odds Ratio<sup>a</sup><br>(95% CI) | No. (%) of COVID-19 related death |  | Odds Ratio<sup>a</sup><br>(95% CI) |
| --- | --- | --- | --- | --- | --- | --- | --- | --- | --- |
|  | Exposed | Unexposed |  | Exposed | Unexposed |  | Exposed | Unexposed |  |
| By sex |  |  |  |  |  |  |  |  |  |
| Female | 177/28797<br>(0.6) | 666/203372<br>(0.3) | 1.28 (1.06, 1.54) | 101/28797<br>(0.4) | 296/203372<br>(0.1) | 1.58 (1.22, 2.04) | 39/28797<br>(0.1) | 81/203372<br>(0.04) | 1.98 (1.29, 3.06) |
| Male | 216/22018<br>(1.0) | 718/166861<br>(0.4) | 1.61 (1.35, 1.91) | 136/22018<br>(0.6) | 398/166861<br>(0.2) | 1.75 (1.41, 2.18) | 66/22018<br>(0.3) | 152/166861<br>(0.1) | 2.07 (1.49, 2.87) |
| By age (by tertiles) |  |  |  |  |  |  |  |  |  |
| ≤64 | 84/17762 (0.5) | 571/129556<br>(0.5) | 0.89 (0.69, 1.14) | 41/17762<br>(0.2) | 226/129556<br>(0.2) | 1.02 (0.71, 1.48) | 12/17762<br>(0.1) | 19/129556<br>(0.01) | 2.17 (0.91, 5.19) |
| 65-72 | 113/15279<br>(0.7) | 269/112645<br>(0.2) | 1.83 (1.42, 2.36) | 76/15279<br>(0.5) | 149/112645<br>(0.1) | 2.00 (1.44, 2.76) | 28/15279<br>(0.2) | 41/112645<br>(0.04) | 2.66 (1.51, 4.67) |
| ≥73 | 196/17774<br>(1.1) | 544/128032<br>(0.4) | 1.80 (1.50, 2.16) | 120/17774<br>(0.7) | 319/128032<br>(0.2) | 1.92 (1.52, 2.42) | 65/17774<br>(0.4) | 173/128032<br>(0.1) | 1.77 (1.29, 2.43) |
| By BMI level, kg/m<sup>2</sup> |  |  |  |  |  |  |  |  |  |
| <18.5 | 1/338 (0.3) | 7/1757 (0.4) | 0.63 (0.06, 6.76) | 1/338 (0.3) | 4/1757 (0.2) | 1.40 (0.10, 19.1) | 0/338(0) | 3/1757 (0.2) | - |
| 18.5-24.9 | 95/14050 (0.7) | 322/124227<br>(0.3) | 1.75 (1.34, 2.29) | 56/14050<br>(0.4) | 132/124227<br>(0.1) | 2.24 (1.54, 3.25) | 22/14050<br>(0.2) | 38/124227<br>(0.03) | 2.44 (1.30, 4.56) |
| 25.0-29.9 | 143/20279<br>(0.7) | 597/157827<br>(0.4) | 1.42 (1.16, 1.74) | 84/20279<br>(0.4) | 309/157827<br>(0.2) | 1.60 (1.22, 2.09) | 37/20279<br>(0.2) | 98/157827<br>(0.1) | 1.97 (1.29, 3.02) |
| ≥30 | 154/16148<br>(1.0) | 458/86422<br>(0.5) | 1.33 (1.08, 1.63) | 96/16148<br>(0.6) | 249/86422<br>(0.3) | 1.52 (1.17, 1.97) | 46/16148<br>(0.3) | 94/86422 (0.1) | 1.96 (1.32, 2.89) |
| **By smoking status** |  |  |  |  |  |  |  |  |  |
| Never | 141/17913<br>(0.8) | 713/214993<br>(0.3) | 1.66 (1.37, 2.01) | 82/17913<br>(0.4) | 339/214993<br>(0.2) | 1.88 (1.45, 2.42) | 38/17913<br>(0.2) | 89/214993<br>(0.04) | 3.13 (2.09, 4.69) |
| Ever | 244/32482<br>(0.8) | 659/153204<br>(0.4) | 1.23 (1.05, 1.44) | 149/32333<br>(0.5) | 349/153204<br>(0.2) | 1.43 (1.16, 1.76) | 66/32482<br>(0.2) | 140/153204<br>(0.1) | 1.63 (1.19, 2.23) |
| **By the number of somatic comorbidities** |  |  |  |  |  |  |  |  |  |
| 0 | 129/26306<br>(0.5) | 852/291701<br>(0.3) | 1.62 (1.33, 1.96) | 61/26306<br>(0.2) | 393/291701<br>(0.1) | 1.65 (1.25, 2.19) | 26/26306<br>(0.1) | 106/291701<br>(0.04) | 2.38 (1.51, 3.75) |
| 1 | 105/14924<br>(0.7) | 304/59153<br>(0.5) | 1.35 (1.06, 1.71) | 72/14924<br>(0.5) | 155/59153<br>(0.3) | 2.04 (1.51, 2.75) | 27/14924<br>(0.2) | 69/59153 (0.1) | 1.90 (1.18, 3.04) |
| 2 | 89/6393 (1.4) | 142/14758<br>(1.0) | 1.67 (1.25, 2.23) | 56/6393 (0.9) | 88/14758 (0.6) | 1.78 (1.24, 2.55) | 26/6393 (0.4) | 33/14758 (0.2) | 2.15 (1.23, 3.75) |
| ≥3 | 70/3192 (2.2) | 86/4621 (1.9) | 1.29 (0.91, 1.84) | 48/3192 (1.5) | 58/4621 (1.2) | 1.23 (0.80, 1.89) | 26/3192 (0.8) | 25/4621 (0.5) | 1.74 (0.94, 3.20) |
| **By Townsend deprivation score** |  |  |  |  |  |  |  |  |  |
| Low | 78/12794 (0.6) | 355/127557<br>(0.3) | 1.67 (1.28, 2.18) | 51/12794<br>(0.4) | 169/127557<br>(0.1) | 2.23 (1.58, 3.16) | 23/12794<br>(0.2) | 62/127557<br>(0.05) | 2.20 (1.29, 3.72) |
| Middle | 108/14886<br>(0.7) | 432/125426<br>(0.3) | 1.52 (1.20, 1.91) | 63/14886<br>(0.4) | 227/125426<br>(0.2) | 1.57 (1.15, 2.15) | 25/14886<br>(0.2) | 68/125426<br>(0.1) | 2.09 (1.25, 3.47) |
| High | 297/23135 | 597/117250 | 1.35 (1.13, 1.61) | 123/23135 | 298/117250 | 1.62 (1.27, 2.05) | 57/23135 | 103/117250 | 1.95 (1.36, 2.82) |

|  | (0.9) | (0.5) |  | (0.5) | (0.2) |  | (0.2) | (0.1) |  |
| --- | --- | --- | --- | --- | --- | --- | --- | --- | --- |
| **By time periods** |  |  |  |  |  |  |  |  |  |
| Before March<br>31<sup>st</sup> | 82/50815 (0.2) | 258/370233<br>(0.1) | 1.68 (1.27, 2.22) | 76/50815<br>(0.1) | 216/370233<br>(0.1) | 1.82 (1.36, 2.43) | 22/50815<br>(0.04) | 53/370233<br>(0.01) | 1.96 (1.12, 3.42) |
| April 1<sup>st</sup> to May<br>31<sup>st</sup> | 275/50733<br>(0.5) | 990/369975<br>(0.3) | 1.40 (1.20, 1.63) | 145/50739<br>(0.3) | 451/370017<br>(0.1) | 1.55 (1.25, 1.91) | 83/50793<br>(0.2) | 180/370180<br>(0.05) | 2.04 (1.52, 2.74) |
| June 1<sup>st</sup> onward | 36/50458 (0.1) | 136/368985<br>(0.04) | 1.31 (0.87, 1.97) | 16/50594<br>(0.04) | 27/369566<br>(0.01) | 2.6 (1.30, 5.19) | 0/50710 (0) | 0/370000 (0) | - |

<sup>a</sup> Logistic regression models were adjusted for birth year, sex, race/ethnicity, body mass index, smoking status, Townsend deprivation index, and history of chronic cardiac disease, diabetes, chronic pulmonary disease, chronic kidney disease, and asthma.

While there was no clear trend regarding time since the first diagnosis of pre-pandemic psychiatric disorder, the relative risk of COVID-19 increased with the number of pre-pandemic psychiatric disorders (Table 3). For example, the OR for all COVID-19 was 1.30 (95%CI 1.13 to 1.50) for individuals with one pre-pandemic psychiatric disorder, 1.84 (95%CI 1.49 to 2.28) for patients with two, and 2.15 (95%CI 1.49 to 3.10) for patients with more than three psychiatric disorders *(p* for trend <0.001).

**Table 3.** Risk of COVID-19 among individuals with pre-pandemic psychiatric disorders, by different psychiatric indicators, compared to individuals without

|  | No. (%) of all COVID-19 |  | Odds Ratio<sup>a</sup><br>(95% CI) | No. (%) of inpatient COVID-19 |  | Odds Ratio<sup>a</sup><br>(95% CI) | No. (%) of COVID-19 related death |  | Odds Ratio<sup>a</sup><br>(95% CI) |
| --- | --- | --- | --- | --- | --- | --- | --- | --- | --- |
|  | Exposed | Unexposed |  | Exposed | Unexposed |  | Exposed | Unexposed |  |
| Time since the first diagnosis of psychiatric disorders, years |  |  |  |  |  |  |  |  |  |
| <1 | 53/5212 (1.0) |  | 2.05 (1.55, 2.72) | 35/5212 (0.7) |  | 2.59 (1.83, 3.66) | 8/5212 (0.2) |  | 1.57 (0.77, 3.19) |
| 1-2 | 74/9693 (0.8) |  | 1.54 (1.21, 1.95) | 38/9693 (0.4) |  | 1.52 (1.09, 2.12) | 23/9693 (0.2) |  | 2.48 (1.59, 3.84) |
| 3-4 | 59/8778 (0.7) | 1384/370233 | 1.31 (1.00, 1.71) | 33/8778 (0.4) | 694/370233 | 1.41 (0.99, 2.02) | 19/8778 (0.2) | 233/370233 | 2.18 (1.34, 3.54) |
| 5-9 | 132/18648 (0.7) | (0.4) | 1.29 (1.06, 1.56) | 81/18648 (0.4) | (0.2) | 1.53 (1.19, 1.96) | 34/18648 (0.2) | (0.1) | 1.71 (1.16, 2.53) |
| ≥10 | 75/8484 (0.9) |  | 1.41 (1.11, 1.80) | 50/8484 (0.6) |  | 1.82 (1.35, 2.46) | 21/8484 (0.2) |  | 2.31 (1.45, 3.69) |
| The number of pre-pandemic psychiatric disorders |  |  |  |  |  |  |  |  |  |
| 1 | 261/39193 (0.7) |  | 1.30 (1.13, 1.50) | 162/39193 (0.4) |  | 1.55 (1.28, 1.89) | 69/39193 (0.2) |  | 1.73 (1.29, 2.32) |
| 2 | 100/9422 (1.1) | 1384/370233 | 1.84 (1.49, 2.28) | 57/9422 (0.6) | 694/370233 | 2.04 (1.53, 2.71) | 30/9422 (0.3) | 233/370233 | 3.25 (2.17, 4.87) |
| ≥3 | 32/2200 (1.5) | (0.4) | 2.15 (1.49, 3.10) | 18/2200 (0.8) | (0.2) | 2.40 (1.47, 3.91) | 6/2200 (0.3) | (0.1) | 2.60 (1.13, 6.03) |

<sup>a</sup> Logistic regression models were adjusted for birth year, sex, race/ethnicity, body mass index, smoking status, Townsend deprivation index, and history of chronic cardiac disease, diabetes, chronic pulmonary disease, chronic kidney disease, and asthma.

Ascertainment of pre-pandemic psychiatric disorders based on primary diagnoses in the UK Biobank inpatient hospital data revealed largely identical results (Supplementary Table 2). Also, restricting to the study period before 27^th^ April 2020 did not modify these estimates (Supplementary Table 3).

## Discussion

Based on a large-scale community-based cohort in the UK, we found that individuals with pre-pandemic psychiatric disorders were at elevated risk of COVID-19, particularly of COVID-19 related hospitalizations and mortality. These associations were independent of many potential confounders, such as socioeconomic status, smoking, and history of somatic comorbidities, and were stronger among individuals with multiple pre-pandemic psychiatric disorders. Notably, we also observed similar increase in the risk of other infections that required hospitalizations during the COVID-19 outbreak in this vulnerable population, supporting the hypothesis that psychiatric disorders may alter the people’s susceptibility to COVID-19 through compromised immunity.

Although with few comparable data in the setting of COVID-19, the association between psychiatric illness and subsequent infections has been consistently demonstrated in previous studies. A number of experimental studies suggested a dose-dependent association between psychological stress and acute infectious respiratory illness. Similarly, using large population-based registers, studies - have shown the association between psychiatric disorders, especially stress-related disorder, and subsequent severe infections in the general population. One recent study conducted in the UK Biobank population^10^ finally brought this picture to COVID-19 scenario, indicating that a range of psychosocial factors, including self-reported symptoms of psychological distress, was associated with increased risk of COVID-19 hospitalization. Clinically confirmed psychiatric disorders were though not addressed in this study. Our new findings of the association between clinically confirmed psychiatric disorders and markedly increase risk of COVID-19, especially severe and fatal COVID-19, underscore the need of surveillance and care in vulnerable populations with history of psychiatric disorders during the COVID-19 outbreak.

Although the underlying mechanisms for the documented asscoiation remain unclear, the activation of hypothalamic-pituitary-adrenal axis among individuals with adverse psychiatric conditions has been widely reported^7 26^. This can lead to altered circulating glucocorticoids and subsequently suppressed cell-mediated and humoral immunity^28^. As a result, the weakened immune system may increase people’s risk of being infected by SARS-CoV-2, when exposed. Furthermore, the overproduction of inflammatory cytokines induced by glucocorticoid receptor resistance^29 30^ can play an important role in the progression of severe infections. Indeed, the correlation of increased cytokine levels with disease deterioration and fatal outcomes of COVID-19 has been reported^31^.

Alternatively, given that COVID-19 is a communicable disease, another explanation for the observed association can be, individuals with psychiatric disorders may have different mobility pattern or suboptimal capacity for mitigating strategies (i.e., social distancing, mask wearing, personal hygiene) during the COVID-19 outbreak, compared with the general population. For instance, it is possible that this population may experience difficulties in maintaining social distancing, or to follow the mobility restrictions^32^, which might consequently increase their possibilities of being exposed to COVID-19 cases. However, our finding of increased susceptibility to other severe infections, which are mainly non-communicable and therefore irrelevant to any mitigating strategies or mobility patterns, among this population argues against a predominant influence of such behavior-related factors.

The major merit of our study is the use of longitudinal data where information bias was minimized since the diagnosis and registration of exposure (i.e., psychiatric disorder diagnosed before the outbreak) and outcome (i.e., COVID-19) was compiled prospectively and independently. The application of the positive control outcome, i.e., hospitalization for other infections, supported a pathway between psychiatric disorders and susceptibility to infections in general, shedding light on the underlying mechanism linking pre-pandemic psychiatric disorders to COVID-19 risk. The large study population in the UK Biobank enabled detailed analyses for all subgroups, and the availability of extensive phenotypic data allowed for consideration of a wide range of important confounding factors.

This study has several notable limitations. First, given that individuals with psychiatric disorders may for their established contact with the health care system have more opportunities to report their respiratory symptoms and get tested for COVID-19, surveillance bias may be a concern. Nevertheless, the obvious shortage of medical resource in the UK during the COVID-19 outbreak should have largely reduced the possibility of hospital admission due to mild health conditions. Also, we observed even stronger associations for severe outcomes (i.e., inpatient COVID-19 and COVID-19 related death) throughout the outbreak period, suggesting that the influence of surveillance bias in the reported associations should be limited. Second, because the accessibility to COVID-19 test in the UK was markedly enhanced within the later part of the study period, heterogenicity in the identified COVID-19 cases can be anticipated. This concern can be partly released by similar estimates obtained for the sub-analyses by different time periods, as well as the sensitivity analysis restricting to the study period before April 27^th^. Last, the UK Biobank recruited only 5.5% of the target population. It is not representative of the general population regarding a number of characteristics and therefore requires cautions when used for the assessment of exposure-disease relationships^33^. However, the reproducibility of the observed association for risk factors of various health endpoints, such as cardiovascular disease, cancer, and suicide has been satisfactory^34^.

In conclusion, in the UK Biobank population, pre-pandemic psychiatric disorders were associated with a subsequently elevated risk of COVID-19, especially COVID-19 related hospitalization and mortality. Although requires further investigations, the similar strength of association observed for hospitalization due to other infections suggests the importance of immune pathways.

## Data Availability

Data from UK Biobank (http://www.ukbiobank.ac.uk/) are available to all researchers upon application. Part of this research has been conducted using the UK Biobank Resource under Application 54803.

## Acknowledgments

We thank the team members involved in West China Biomedical Big Data Center for Disease Control and Prevention for their support. Huan Song had full access to all the data in the study and takes responsibility for the integrity of the data and the accuracy of the data analysis.

## Author Contributions

UAV, HS, and FF were responsible for the study concept and design. YH, YS, ZY, and YQ did the data and project management. HY, WC, YC, YZ, JH did the data cleaning and analysis. HY, WC, YC, YZ, DL, FF, UAV, and HS interpreted the data. HY, WC, DL, FF, UAV, and HS drafted the manuscript. All authors approved the final manuscript as submitted and agree to be accountable for all aspects of the work.

## Ethical approval

UK Biobank has full ethical approval from the NHS National Research Ethics Service (reference number: 16/NW/0274), and the study was also approved by the biomedical research ethics committee of West China Hospital (reference number: 2020.661).

## Transparency

The study guarantor (HS) affirm that this manuscript is an honest, accurate, and transparent account of the study being reported; that no important aspects of the study have been omitted; and that any discrepancies from the study as planned (and, if relevant, registered) have been explained.

## Copyright statement

This is an Open Access article distributed in accordance with the Creative Commons Attribution Non Commercial (CC BY-NC 4.0) license, which permits others to distribute, remix, adapt, build upon this work non-commercially, and license their derivative works on different terms, provided the original work is properly cited and the use is non-commercial. See: http://creativecommons.org/licenses/by-nc/4.0/.

